# “The new gay plague”: analysis of public attitudes toward monkeypox

**DOI:** 10.1101/2022.11.01.22281797

**Authors:** Tej Shah

## Abstract

Monkeypox was declared a public health emergency on August 4, 2022, in the United States. The emerging isolation of the virus in the LGBTQ+ community—particularly among gay, bisexual, and men who have sex with men (GBMSM)—has led many to draw parallels between the emerging multi-country outbreak and the 1980s HIV/AIDS crisis. The purpose of this study was to investigate media framing of the monkeypox outbreak in American media through the lens of HIV social constructionist theory. Content analysis of a sample of 59 articles from the top-five most viewed U.S. media outlets was collated against quantitative trends in word frequencies in monkeypox-related tweets (n = 255,363). Results found that coverage often framed monkeypox as a product of GBMSM hypersexuality, leading to potentially stigmatizing perceptions and the drastic increase in tweet content related to sexual activity. While greater attention to stigma was observed in coverage, blame attribution to populations, governments, and practices was one of the most common frames across all media sources. Heavy reporting of systemic barriers to vaccination, testing, or diagnosis serve as continuities from HIV/AIDS and COVID-19 epidemics, underscoring fears around a second plague and influencing public attitudes. Monkeypox conspiracy theories also proliferated heavily on Twitter, with a noticeable increase in conspiracy language over time. These findings can inform the social realities of monkeypox, an understudied dimension, of which an understanding is vital to implementing services that address all elements of the ongoing outbreak.

## Introduction

On May 7, 2022, the National IHR Focal Point for the United Kingdom notified the World Health Organization (WHO) of a confirmed case of human monkeypox (MPX) in an individual who had traveled from the United Kingdom to Nigeria (CDC, 2022a). While the virus has been endemic in Central and West African regions since 1970 (Kozlov, 2022), it has rarely occurred outside of the African Continent. After the U.K case was identified, however, MPX quickly spread through non-endemic areas, including Europe and North America (Thornhill et al., 2022). As of October 2022, there have been over 69,244 confirmed cases across over 107 countries, the majority of which are historically non-endemic (CDC, 2022a). While the World Health Organization has declared the multi-country outbreak to be an “public health emergency of international concern” (WHO, 2022), the novel, disproportionate spread of the virus among gay, bisexual, and other men who have sex with men (GBMSM)—over 98% of total cases in the United States (Thornhill et al., 2022)—has engendered potentially stigmatizing, misinformative rhetoric from news media sites and the general public alike (Chang et al., 2022). This distinct prevalence in the LGBTQ+ community has caused many to draw parallels between the current MPX crisis and 1980s HIV/AIDS epidemic (Diamond, 2022). While these two viruses are hardly alike, the influential role of media in shaping perceptions and attitudes towards both MPX and HIV is evident.

This paper will examine the public attitudes and perceptions toward monkeypox (MPX) in the United States, analyzing media coverage and relevant tweets through the lens of HIV signification theory and assessing implications for health disparities in the continuities and changes between the two health crises. The focus will be on the social and technological relationships which exacerbate misinformation, stigmatization of MSM, and barriers to treatment. These findings may be helpful to health professionals interested in understanding how to better address monkeypox-related stigma, and how to reduce barriers to interventions like vaccination or testing. First, this paper will begin with a clinical review of existing literature on monkeypox, followed by a background on social constructionism, media frames, and their connections to the 1980s HIV/AIDS crisis in the United States. Subsequent sections will describe the research methodology, results, data analysis, limitations, and future recommendations.

### Epidemiology of Monkeypox

Monkeypox virus (MPXV), a double-stranded DNA virus belonging to the genus Orthopoxvirus, is a re-emerging zoonotic pathogen known to cause dermatological lesions or lymphadenopathy in humans (McCollum & Damon, 2014). Human monkeypox (MPX) was first reported in 1970 in the Democratic Republic of the Congo (Kozlov, 2022); following the eradication of smallpox in the 1980s, mass vaccination was ceased despite it offering some immunity against monkeypox (Islam et al., 2022). Since then, the infection has been endemic in multiple Central and West African countries, where the virus has been isolated in several species of small mammals and non-human primates (World Health Organization, 2018). Human-to-human transmission is believed to occur through direct contact with infectious lesions on the skin, as well as inhalation of respiratory droplets or indirectly via fomites like bedding, clothing, or other surfaces (McCollum & Damon, 2014).

Historically, patients with monkeypox have presented with fever, lymphadenopathy, head or muscle aches, respiratory symptoms, and the development of maculopapular or vesiculopustular rash (McCollum & Damon, 2014). While there are no vaccines formulated specifically against monkeypox, the use of the JYNNEOS smallpox vaccine has been approved by the FDA during the current multi-country outbreak given its 85% efficacy rate (Center for Biologics Evaluation and Research, 2021). On June 28, 2022, the United States federal government announced the rollout of the JYNNEOS vaccine in limited supply and has relied on local health departments, health-based nonprofits, hospital centers, and other public health agencies to manage the distribution of the vaccines (New Mexico Department of Health, 2022). Current eligibility for vaccination varies on local supply but tends to include gay, bisexual, and men who have sex with men (GBMSM) with multiple sexual partners, other LGBTQ+ individuals aged 18 years or older, sex workers, individuals working with food, and close contacts for anyone engaging in possible risk behaviors (CDC, 2022a).

### HIV/AIDS, Media Frames, and Meaning Construction

In June of 1981, the CDC published an article describing rare cases of *Pneumocystis carinii pneumonia* (PCP) in five previously healthy gay men in Los Angeles (McKay, 2017). As cases grew, an overwhelming majority were reported among young men who identified as gay or bisexual or had reported having sex with another male prior to hospitalization; therefore, the mysterious illness was named GRID: gay-related immune deficiency (Platt & Platt, 2013). In the time it took for former President Ronald Reagan to publicly address the disease, a variety of inflammatory meanings had risen out of hysteria and spread among the American population in parallel. Historical research by Treichler (1987) compiled meanings that include, but are not limited to:

1. A gay plague, probably emanating from San Francisco
2. A Soviet plot to destroy capitalists
3. A plague stored in King Tut’s tomb and unleashed when the Tut exhibit toured the US in 1976
4. A creation of biomedical scientists and the CDC to generate funding for their activities
5. A fascist plot to destroy homosexuals
6. God’s punishment of our weaknesses
7. The price paid for anal intercourse
8. A disease that turns fruits into vegetables
9. A disease introduced by aliens to weaken us before the takeover
10. Nature’s way of cleaning house

This accumulation of social constructions ranging from mystical to purely erroneous not only worsened the burden of disease (Heller, 2015), but contributed to rapidly increasing stigmatization of GBMSM (Jeffries et al., 2015). Many stemming from sensational media reporting, Myrick (1998) analyzed the construction of HIV/AIDS and gay identity in two major Southern newspapers during the U.S. AIDS crisis, finding that both papers marginalized gay sexualities and ultimately silenced queer voices (Myrick, 1998). In fact, despite the importance of news media in HIV/AIDS education (Backstrom & Robins, 1998; Brodie et al., 2004), existing literature suggests that reductive media framing and erasure worsened disparities in race and sexual orientation (Cohen, 2009; Levenson, 2005; Persson & Newman, 2008).

Sociologists, public health officials, and HIV scholars often engage the idea of media frames in the contexts of social constructionism and the foundation of reality (Herek et al., 2003; McCauley et al., 2013; Tierney et al., 2006). The way an idea is framed, much like language at large, can simultaneously highlight its specific elements while hiding others (Gamson et al., 1992). As such, media forms i.e. news content, do not simply reflect reality: they define it (Secrist, 1986). The discursive use of metaphors, narrative maneuvers, or personal bias play significant roles in shaping collective knowledge, leaning into the interpretation of the world around us (Yan, 2020). As our world is revolutionized by globalization and technological progress, more and more as individuals we “walk around with media-generated images of the world, using them to construct meaning about political and social issues” (Gamson et al., 1992).

Many sociologists and medical anthropologists have argued that media framings of illness are embedded in certain sociocultural contexts (Sherlaw & Raude, 2013; Staniland & Smith, 2013; Washer, 2004). Most often, a distinction is made between disease, the biological condition, and illness, the social connotation of this disease (Eisenberg, 1977). These sociocultural dimensions play important roles in determining how illness is experienced, addressed with policy, and, most importantly, characterized by social perceptions and attitudes (Conrad & Barker, 2010). For example, Sastry and Dutta (2011) found that third-world or childlike U.S. media representations of HIV/AIDS in India furthered postcolonialist perceptions of India and the global East (Sastry & Dutta, 2011). Across the dearth of public health crises covered by media from mental illness to infectious disease (Gui, 2021; 2018; Sieff, 2003; Tierney et al., 2006), it is well recognized that media possess power as interpretive vehicles of culture (Spitulnik, 1993). In the highly digitized and globalized environment of the United States, the framing of illness can create public anxiety and even dictate public health policy (Saksena, 2018).

The increasing prevalence of social media is changing the ways in which information on world issues is gathered, disseminated, and discussed. In the United States, more than half of U.S. adults report getting news via social media “often” or “sometimes” (Liedke & Matsa, 2022). Twitter is one of the most popular social media and microblogging sites. In 2022, over 500 million tweets were posted per day (Sayce, 2022). Because of the unique digital engagements between celebrities, “semi-public” individuals, corporations, governments, NGOs, and ordinary individuals, Twitter is thought to represent the “full spectrum of communications from personal and private to ‘masspersonal’ to traditional mass media” (Wu et al., 2011). Literature demonstrates that tweet content specifically can be used to analyze public health topics, namely public opinion on COVID-19 (Boon-Itt & Skunkan, 2020) and HPV vaccination (Surian et al., 2016). Because modern media curates and filters content based on users’ engagement history and opinions (Cinelli et al., 2021), individuals develop attitudes about the cognitive and implied attributes of the information presented to them (Messing & Westwood, 2022). This leads to construction of both objective and subjective reality, forging a new perception of “objective” truth (Wohn & Bowe, 2014). Evidently, Twitter can provide a great data source for discerning public attitudes and social constructions related to infectious disease.

## Methods

This paper employs a sequential explanatory mixed-method design to analyze qualitative content analyses of news articles pertaining to attitudes around human monkeypox (MPX) in the United States and quantitative data from relevant tweets (n = 255,363). Ethical approval was not necessary as the sample of tweets used in the analysis are publicly available information posted on social media, available via the Harvard Dataverse project.

News articles were selected using the top five most visited sites in July 2022: New York Times, CNN, Fox News, MSN, and Google News (Similarweb, 2022); however, since Google News is a media aggregator and does not publish any independent coverage, it was removed from the list and replaced with New York Post—sixth on the list. Headlines and paragraphs were searched using four different combinations: monkeypox *and* the United States *and* MSM, monkeypox *and* United States *and* men who have sex with men, monkeypox *and* United States *and* gay, and monkeypox *and* the United States *and* LGBTQ. This search strategy yielded 80 articles. Secondary filtering was done to exclude results that were solely video content, did not focus on the United States, or briefly mentioned monkeypox. After the second round of selection, 59 articles remained. Dedoose was used to perform the inductive content analysis, generate a coding scheme, and record overlying themes.

For quantitative analysis, an open source approach is presented in which tweets were collected from the Harvard Dataverse (Thakur, 2022), and then analyzed and visualized using R programming. Informed by the qualitative findings, a list of 100 codewords was generated using keyword network technique (Ávila-Toscano et al., 2018). Keywords were searched and filtered within tweets to analyze trends over time.

### Qualitative Findings

A total of 59 articles were identified between July 1, 2022 and August 8, 2022 after screening and sorting from the 5 online newspapers (CNN: 18, New York Times: 18, MSN: 9, Fox News: 7, New York Post: 6). In terms of political bias, three sources (CNN, NYT, and MSN) were identified as political centrist and left-leaning, while two sources (NYP and FOX) were identified as right-leaning (Allsides, 2019). New York Times had the most monthly viewers at 458.7 million in July 2022, while the New York Post had the least at 163.9 million; all other selected sites fell within this range.

The top three most prominent topics were vaccination (n = 49, 85.9%), MSM (n = 46, 77.96%), and inequities (n = 36, 61.01%). (See **Table 1** for examples of content findings.) The blame frame was the fourth most common frame (n = 34, 57.62%) in American news, summarized in **Table 2**. Additionally, failure framing found itself in almost half of all articles. (n = 28, 47.46%). **Table 3**. and **Table 4**. provide examples of reported barriers to successful vaccination, testing, or implementation or success of any other public health intervention.

**Table 1.** Examples of media content

| Content category | Examples |
| --- | --- |
| Stigmatization of MSM | “Early monkeypox cases spread through close contact at “circuit parties” attended by gay men, but some media outlets, worried about homophobia, shied away from reporting the full truth.”<br>“...the type of close physical contact that occurs at parties where groups of men, often half dressed, dance in close proximity.” |
| Anti-stigma | “Our team is also committed to reducing stigma among the LGBTQ community, which has been singled out and treated unfairly because of this outbreak. No single individual is to blame for the spread of any virus.” |
| Minimization of risk | “The risk to the broader population is small.”<br>“The bug simply isn’t that transmissible, requiring prolonged close contact to spread.” |
| Exaggeration of risk | “Signaling that the virus represents a significant risk to Americans.”<br>“Monkeypox is here to stay.” |
| Comparisons to other diseases | “Experts point to the way HIV spread as a possible indicator for where the virus will go next.” |
| Spill over | “Experts say the disease could spill over into other populations, especially due to vaccine shortages.” |
| Misinformation/bad comparison | “But Dr. Varma wrote that he and other experts fear that monkeypox could now wind up becoming an entrenched sexually transmitted infection like syphilis and H.I.V.”<br>“Regardless of the messaging coming from health officials, monkeypox is a pandemic...” |
| Blame | See **Table 2.** |
| Barriers to vaccination | See **Table 3.** |
| Barriers to diagnosis and surveillance | See **Table 4.** |

**Table 2.** Examples of blame

| Content category | Examples |
| --- | --- |
| Blaming federal government | “The fact is that the US, and the world for that matter, is just bad at responding to infectious disease outbreaks.” |
| Blaming state government | “‘It doesn’t help us to have 57 plans when we have a national challenge,’ she said, referring to the 50 states plus territories and major cities.” |
| Blaming funding | “Monkeypox virus could further spread due to lack of funding to sexual health programs, experts say” |
| Blaming public health, medical system | “‘The CDC has no idea what they’re doing, nobody’s educated (not even doctors) and doctors will refuse to see you,’ she said”<br>“The systems that have been taxed really hard through the coronavirus are the same systems we have to rely on for monkeypox.” |
| Blaming MSM | “Cases in the city are up by 50% since last week, leading some to point fingers at June’s Pride month celebrations.” |
| Blaming COVID-19 | “When added to chronic low staffing, a problem exacerbated by the COVID-19 pandemic, appointments can be hard to come by.” |

**Table 3.** Examples of barriers to vaccination

| Content category | Examples |
| --- | --- |
| Supply as a barrier | “A vaccine supply that has fallen far short of demand” |
| Pandemic fatigue as a barrier | “The public is tired of hearing about infectious disease.”<br>“Two years of pandemic isolation have made people eager for human connection.” |
| Ring vaccination failure as a barrier | “The CDC’s initial strategy “was just doomed to failure... ‘It’s not that we don’t have the contact tracers. It’s that the individuals don’t know their contacts.’” |
| State, federal government as a barrier | “He said people he knows are trying to be careful but are increasingly angry at what they feel is a lack of urgency to protect the gay community in particular...”<br>“We don’t control public health in the 50 states, in the territories and in the tribal jurisdiction... We rely on our partnership to work with them. They need to work with us.” |
| Vaccine hesitancy as a barrier | “And some fear the vaccine itself, in an echo of the hesitancy and mistrust that hindered the coronavirus response.” |
| Technology as a barrier | “Monkeypox Vaccine Rollout is Marred by Glitches in New York”<br>“I think the challenge is that we need to make sure that the vaccine doesn’t go just to the people that are savvy and can get it quickly. We don’t want the vaccine only to go to white wealthy individuals.” |
| Lack of attention as a barrier | ““At the time, there was little resources on finding the vaccine.” |

**Table 4.**
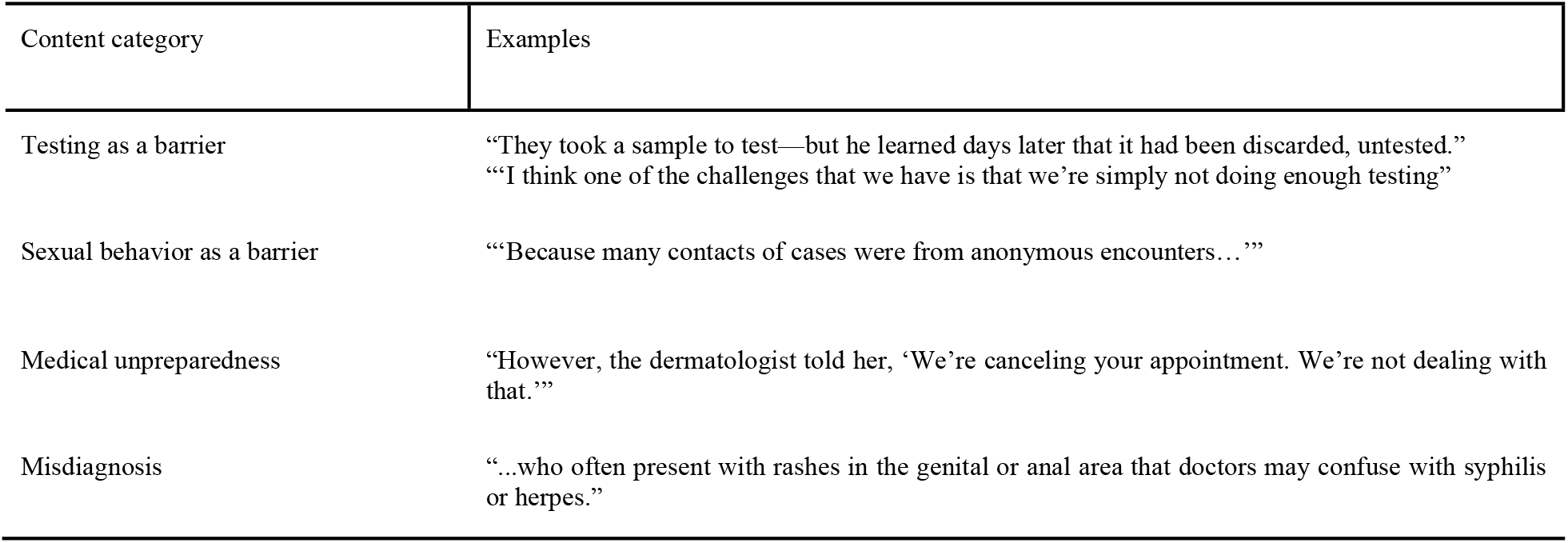
Examples of barriers to diagnosis and surveillance

### Quantitative Findings

A total of 255,363 tweets were quantitatively analyzed from 4 data ranges: May 7, 2022– May 21, 2022 (May), June 5, 2022–June 11, 2022 (Early June), June 12, 2022–June 30, 2022 (Mid to Late June), and July 1, 2022–July 23, 2022 (July). This timeframe represents the months between the first 2022 case of monkeypox and the WHO declaration as a global health emergency. Search terms fell into six categories: LGBTQ+ (n = 15, 14.85%), sexual activity (n = 22, 21.78%), public health (n = 7, 6.93%), conspiracy (n = 33, 32.67%), COVID-19 (n = 7, 6.93%), politics (n = 11, 10.89%), and religion (n = 5, 4.95%).

## Discussion

Mass media has grown to be one of the primary mechanisms through which individuals learn about public health crises and health promotion (Anwar et al., 2020). However, poor framing of these issues can detrimentally influence individuals’ opinions and foster the constructions of misinformative or stigmatizing meanings surrounding them. This paper sought to examine U.S. public attitudes and perceptions towards monkeypox in relation to media framing of the emerging outbreak. Understanding mechanisms of health communication is critical to not only preventing stigma but encouraging the acceptance of new or modified health behaviors.

Content analysis of monkeypox media coverage reveals several clear trends that emerge over the 1-month period spanned by the selected sources. First, inclusivity in public health has increased drastically since the 1980s HIV/AIDS epidemic. Journalists and media providers are paying more attention to stigma and wording choices. Our qualitative findings indicate that greater attention to stigmatization was expressed in almost half of the articles (n = 28, 47.46%). Articles were observed referencing LGBTQ+ representatives or organizations, or even detailing the history of the HIV/AIDS epidemic e.g. “…ACT UP, which formed in 1987 in response to government inaction on H.I.V./AIDS.” Overall, fundamental shifts in collective understanding of health, equity, and LGBTQ+ rights since the 80s were observed driving inclusivity in language and careful consideration of media content. This optimistic finding, in the context of past media behavior during HIV/AIDS (Brodie et al., 2004; Persson & Newman, 2008), illustrates growth since the media-corroborated stigma of the COVID-19 pandemic (Yang et al., 2021).

Despite the progress that has been made, the media response has been far from perfect. The use of contradictory and misinformative comparisons in public media coupled with a lack of accountability, created divergent realities of monkeypox in the United States, many of which are thematically present in tweet data. Writers were seen framing monkeypox simultaneously as something that will ‘spill over’ into the general population and a ‘non-issue,’ as well as comparing the virus to an STI like HIV or syphilis despite statements that it is not an STI. These frames are correlated with an increase in the presence of keywords ‘STI’ and in tweets from the analysis period (Fig 1). These findings are in line with existing research suggesting that competing narratives contribute to polarizing ideas and general confusion (Eliaz & Spiegler, 2020; Kennedy-Hendricks et al., 2016).

**Fig 1.**
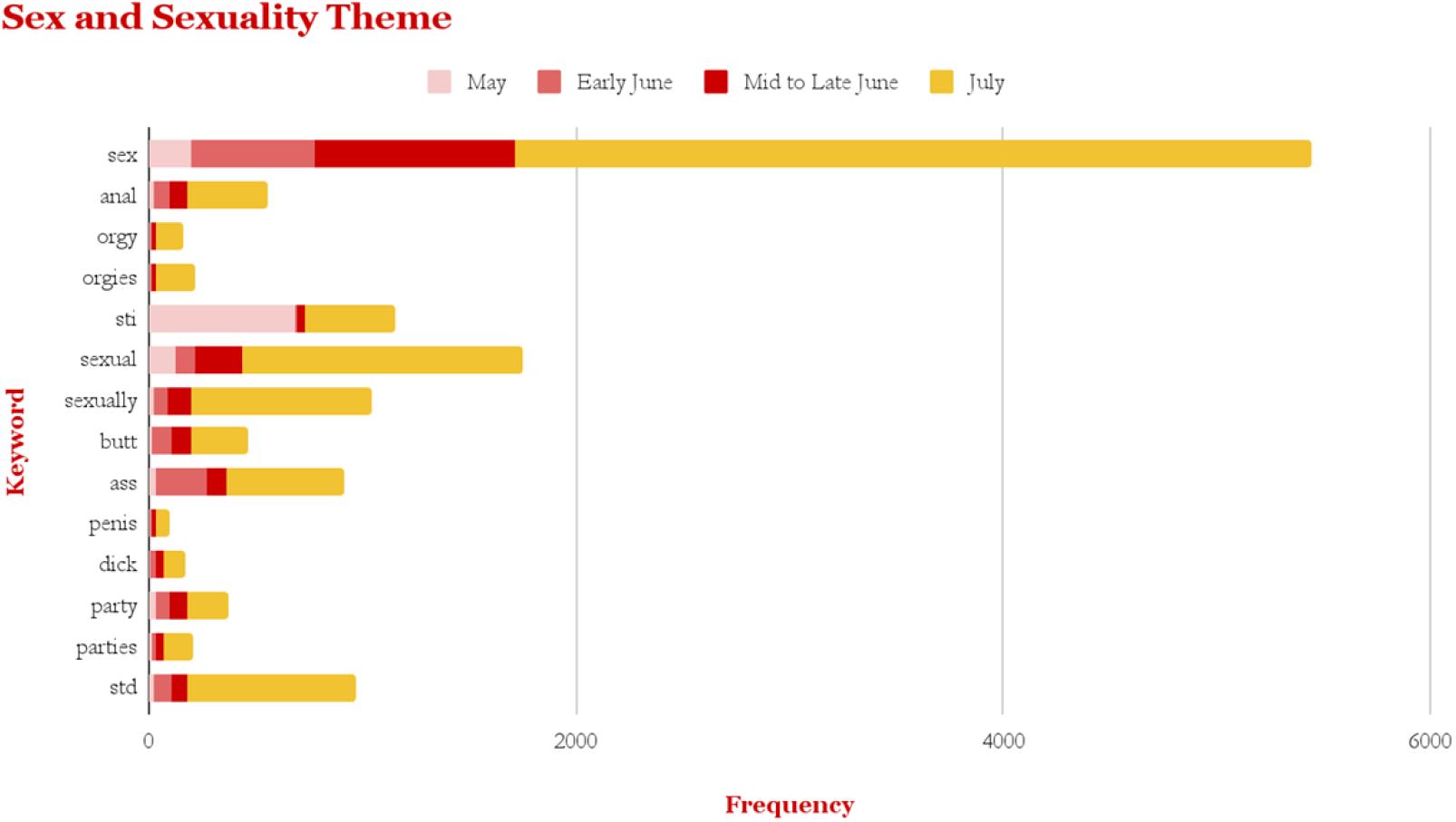
Frequencies of select keywords related to *sex and sexuality* theme from May to July of 2022.

**Fig 2.**
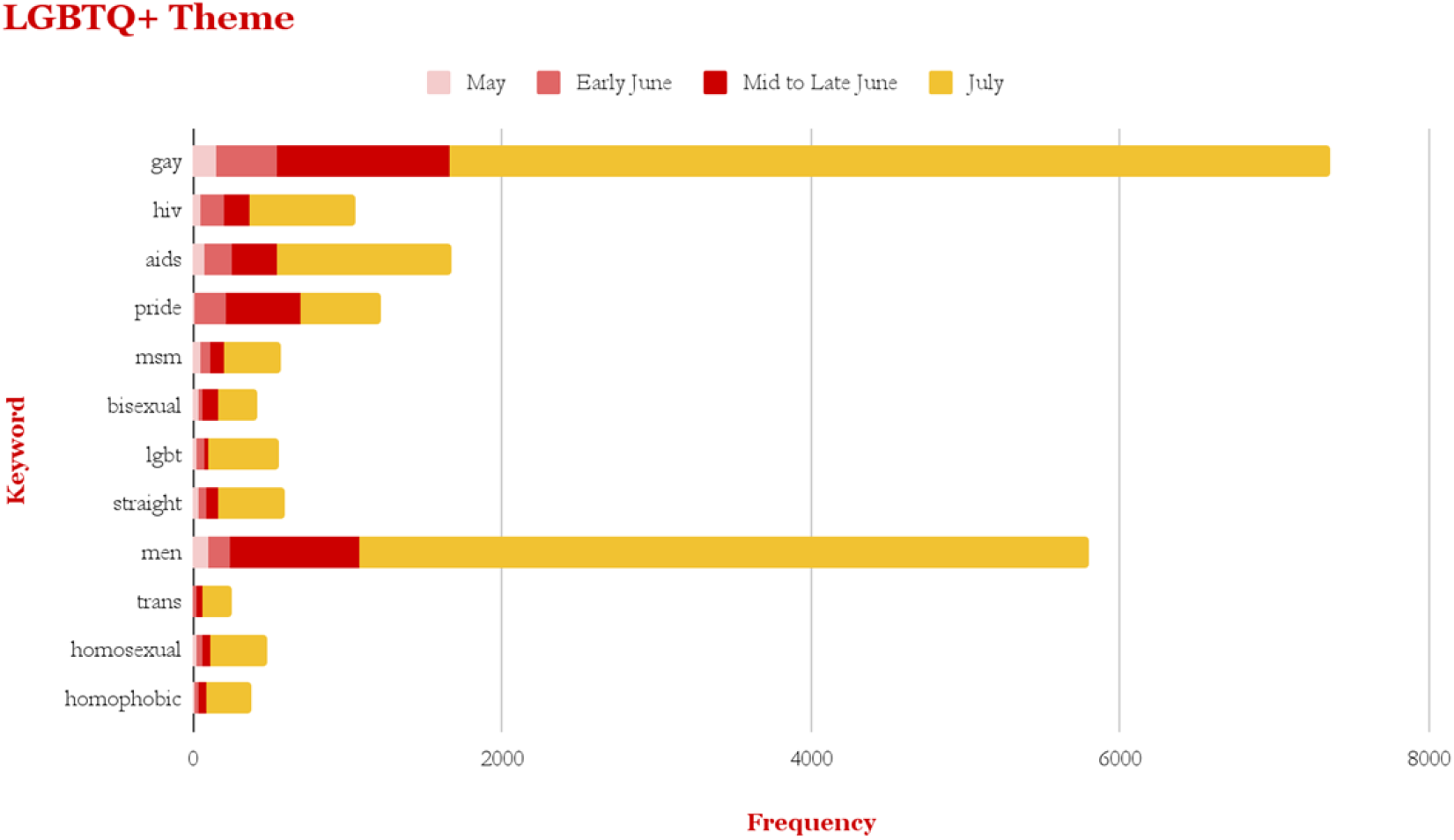
Frequencies of select keywords related to *LGBTQ+* theme from May to July of 2022.

**Fig 3.**
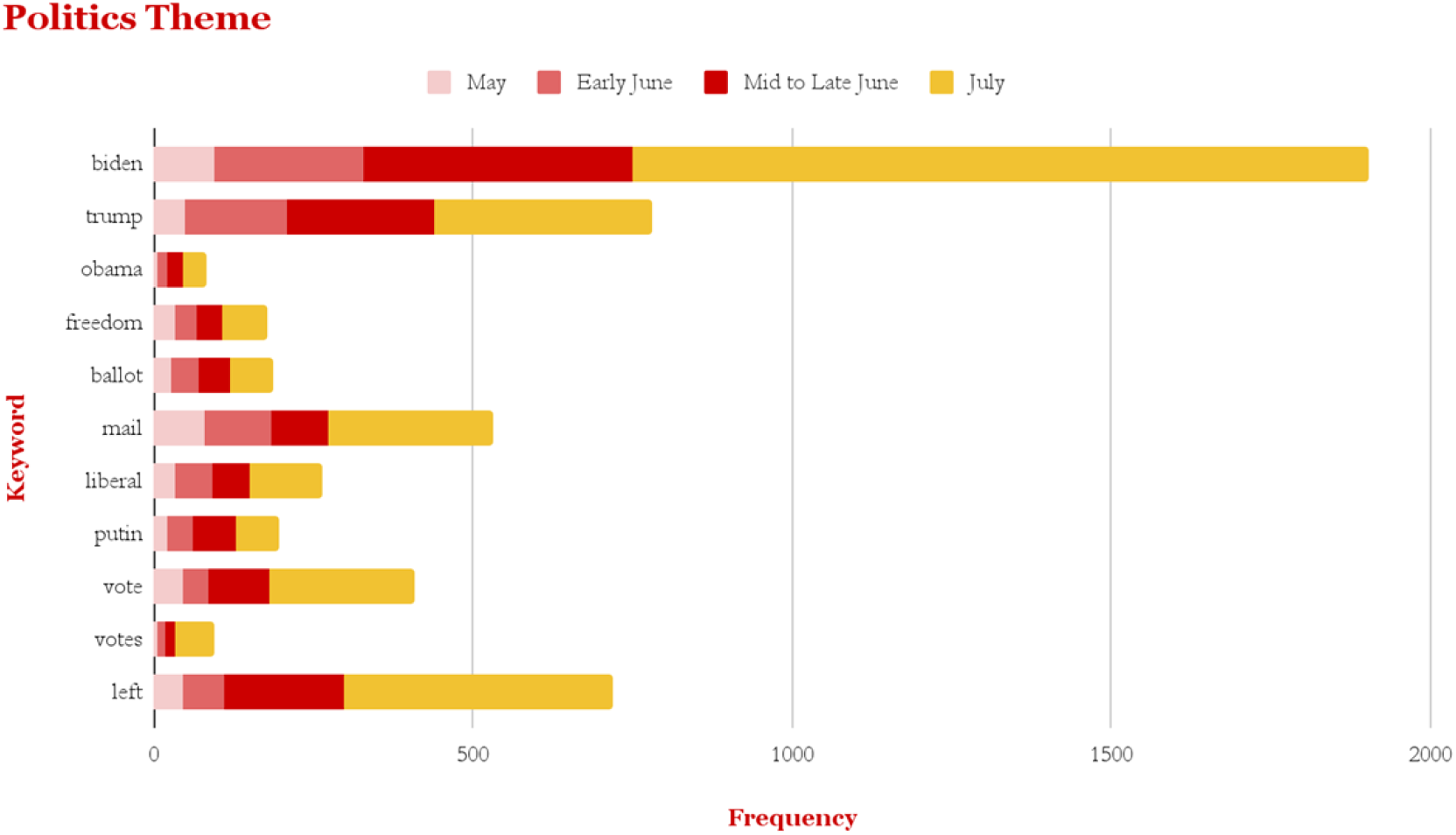
Frequencies of select keywords related to *politics* theme from May to July of 2022.

**Fig 4.**
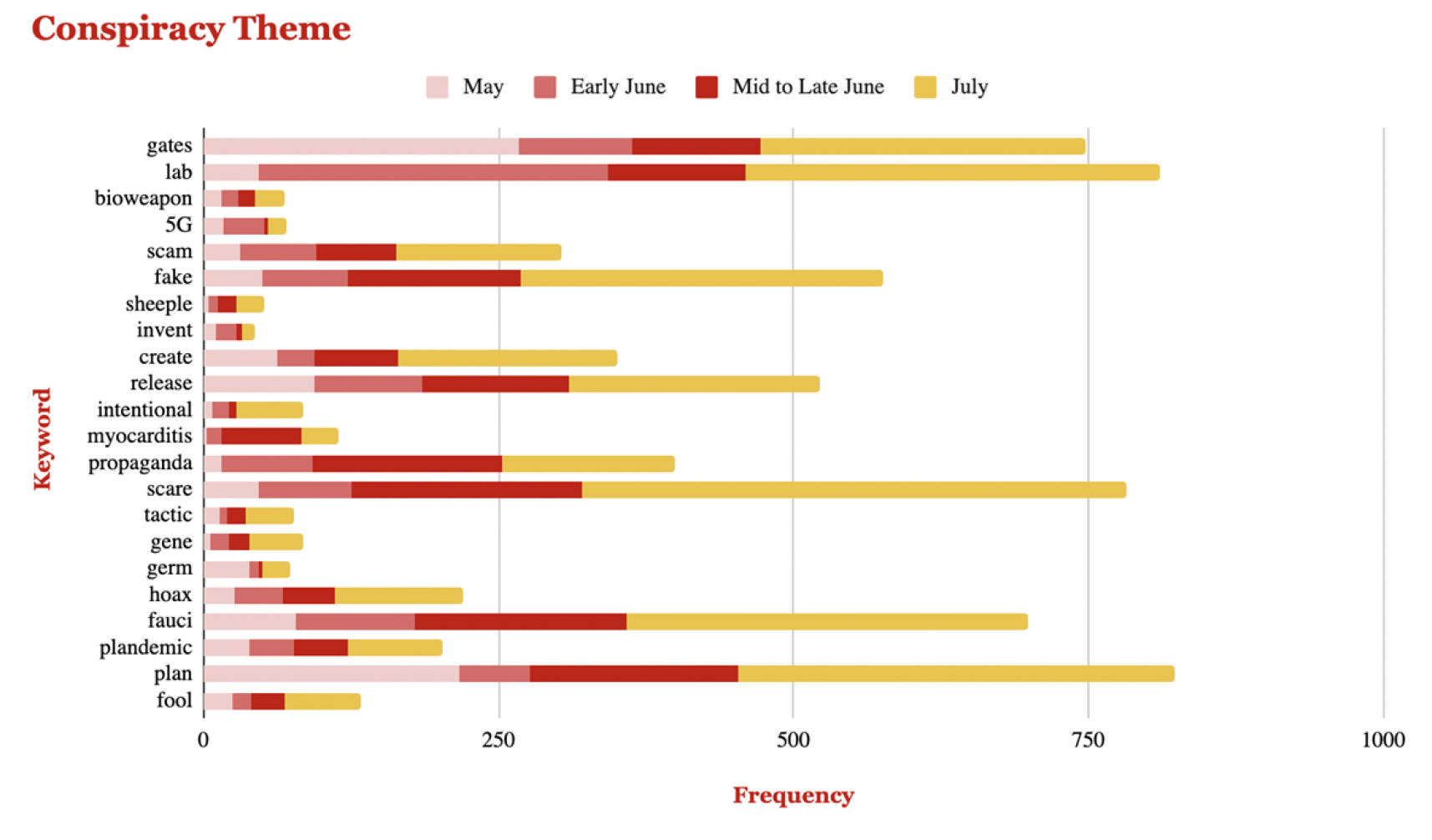
Frequencies of select keywords related to *conspiracy* theme from May to July of 2022.

**Fig 5.**
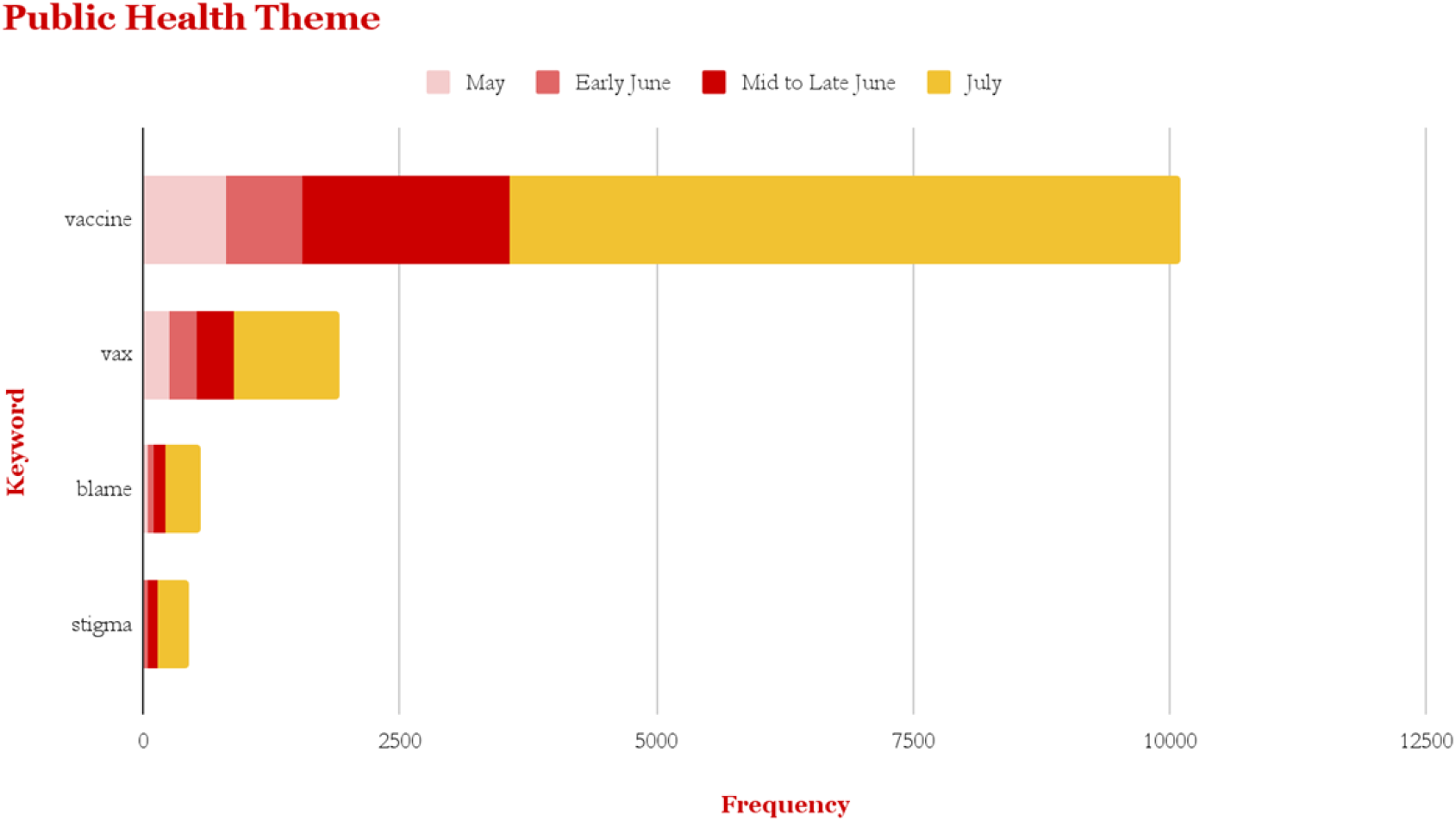
Frequencies of select keywords related to *public health* theme from May to July of 2022.

**Fig 6.**
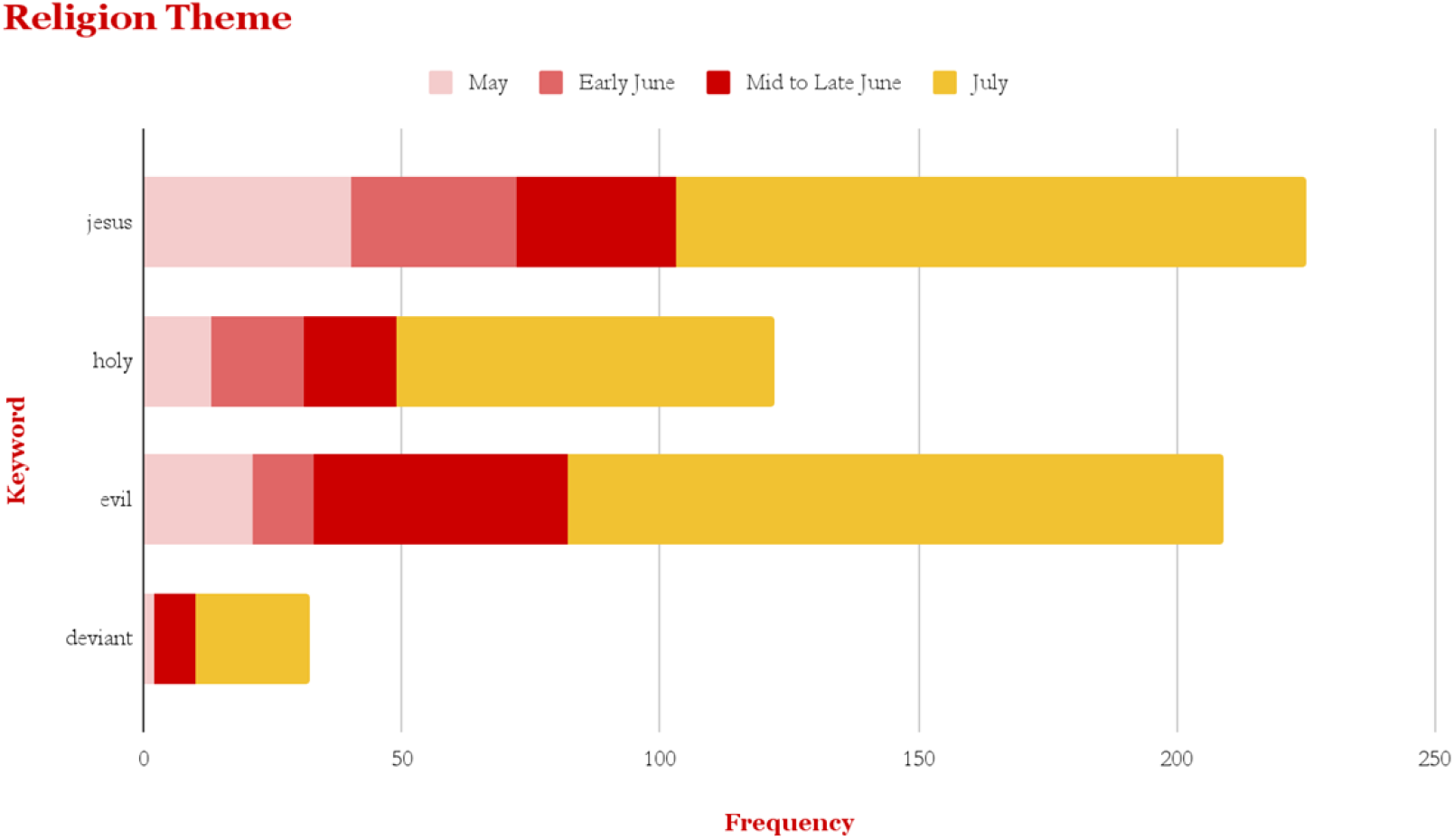
Frequencies of select keywords related to *religion* theme from May to July of 2022.

**Fig 7.**
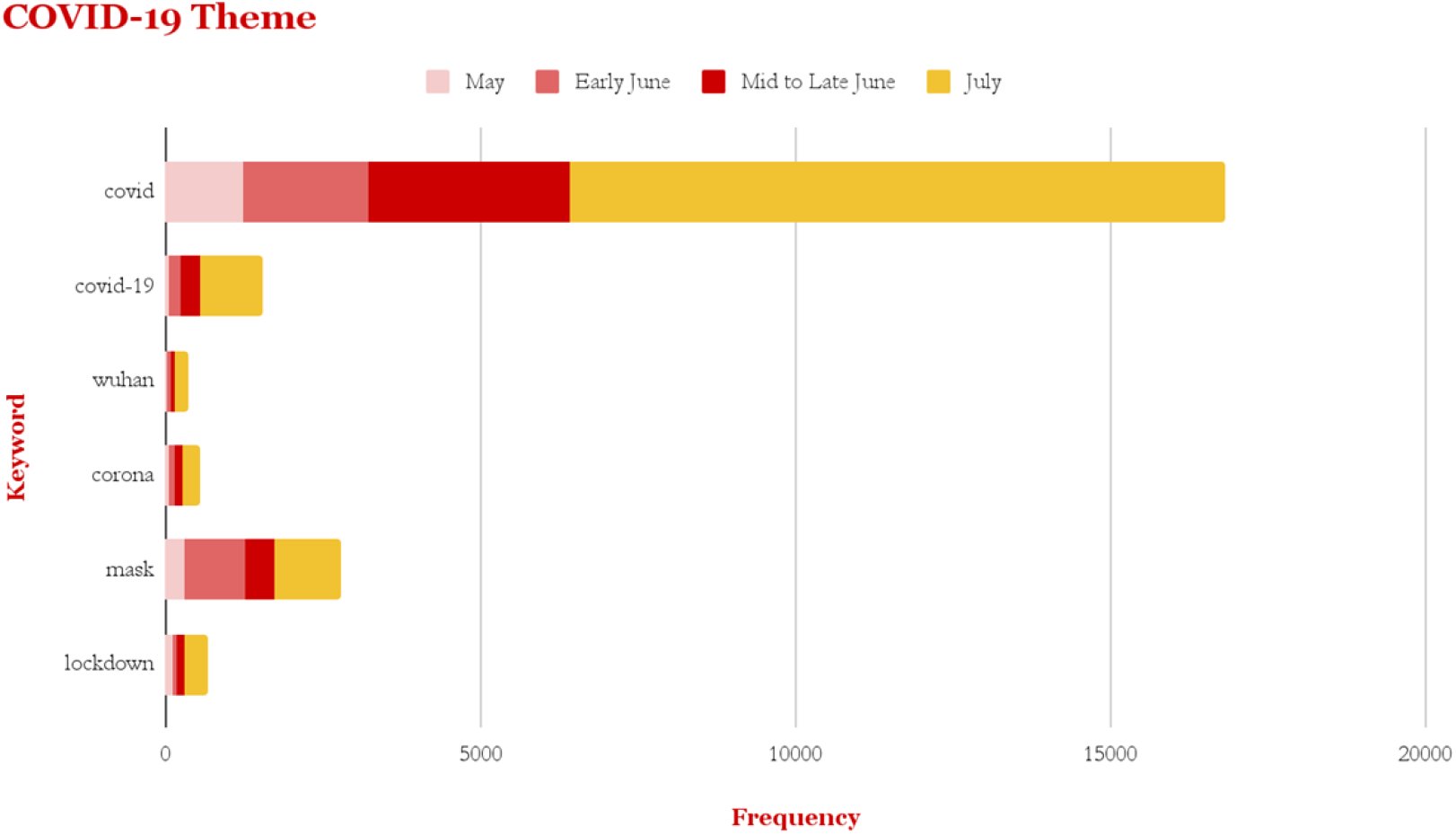
Frequencies of select keywords related to *COVID-19* theme from May to July of 2022.

Additionally, reported failure of the ring vaccination strategy implemented by federal and regional policymakers was often pinned on GBMSM. With success contingent on the vaccination of contacts of confirmed monkeypox patients, public health officials quickly found that anonymous sexual encounters among GBMSM were constraining contact tracing. However, an abundance of existing literature demonstrates that this cultural practice is documented (Garcia et al., 2012). Instead of considering the inappropriateness of ring vaccination for this population, articles tended to associate this failure with clubs of “half dressed men” or “a hot and heavy festival season.” The use of the hypersexual frame could be attributed to the construction of monkeypox meanings that stigmatize behaviors of GBMSM. Descriptive statistics of tweets showed that keywords related to the *sexual activity* theme increased substantially from May to July. For example, the presence of terms ‘orgies,’ ‘anal,’ and ‘ass’ increased by between 1700% and 5900% in the 2 months analyzed. The presence of words like ‘pride’ or ‘party’ also showed increases over time, likely continuing the sexualization of GBMSM culture. The proliferation of these promiscuous associations not only worsen stigmatization (Iott et al., 2022), but operate in contrast to progress in sexual liberation ushered in by HIV/AIDS activists (Schubert, 2022).

Quantitative analysis of tweet content unexpectedly exposed the ongoing proliferation of conspiracy-based misinformation on social media. The presence of keywords ‘gates,’ ‘propaganda,’ ‘plan’ or ‘plandemic,’ ‘hoax,’ ‘fauci,’ and others all increased substantially in the 2 months of data collection. Additionally, *conspiracy* themes were observed to overlap with others. For example, the presence of terms ‘ballot’ or ‘vote’ within the *politics* theme often coincided with the above *conspiracy* keywords; interestingly, users suggested that the emerging monkeypox outbreak was an engineered effort to return to mail-in voting and falsify election outcomes. In this same way, ‘wuhan’ from the *COVID-19* theme often joined *conspiracy* keywords with users insinuating that the same lab that created and released the novel coronavirus released the circulating monkeypox virus. These findings affirm previous research on Twitter misinformation (Jin et al., 2014; Oyeyemi et al., 2014; Sharma et al., 2020), providing new insight into sociodigital patterns relating to infectious disease and the growing need to curb misinformation on digital platforms.

Although many of the comparisons made between HIV and monkeypox were found to be inappropriate e.g. becoming a “…entrenched sexually transmitted infection like syphilis and H.I.V.,” the emerging monkeypox outbreak does mirror HIV on specific points. The substantial use of the inequalities frame, across almost all sources, was generally used in relation to racial disparities with infection, lack of investment into health infrastructure, stigmatizing messaging, and other systemic issues. Analysis found that there is continuity between these systemic issues from the 1980s HIV/AIDS crisis into the current monkeypox outbreak. The media and governmental response to HIV not only gave rise to similar racial disparities (Brier, 2018) as monkeypox is currently demonstrating (CDC, 2022b), but historically manifested almost identically in the healthcare system (Berger, 2022; Piot et al., 2007). With this being stated, monkeypox is still not an extension of HIV: any comparisons to this end remain inappropriate and potentially misinformative. Monkeypox, like COVID-19 and HIV/AIDS, is instead a continual product of a flawed public health system.

Evidence shows there is a relationship between media framing and the construction of social paradigms (Adoni & Mane, 1984). The findings of this study offer insights and opportunities to address public attitudes to not only media providers, but policymakers and public health decision-makers. First, media sources should be aware of the implications of word choice and interpretive issue framing. This capacity may play a role in creating barriers to public health interventions like vaccination, testing, or even general awareness. As such, journalists must ensure that monkeypox frames presented are accurate and understanding, and do not give rise to any personal biases. To the author’s knowledge, this is the first study delving into the social realities of monkeypox in the United States. Greater work by anthropologists, sociologists, and other social scientists delving into meanings of monkeypox illness rather than the solely biological virus is necessary to the development of a clear picture of this ongoing outbreak.

## Limitations

The results of this study are limited by sampling procedures, as only the top five most-viewed sources were analyzed. Smaller news outlets still have the capacity to influence public perceptions of specific issues (King et al., 2017), so this analysis represents a small examination into the vast expanse of online news media coverage of the monkeypox outbreak in the United States. Future studies with wider selection criteria should be done to explore this phenomenon.

Additionally, research by Murshed et al. (2021) found that the use of slang, typos, repeated characters, transposition, concatenated words, complex spelling mistakes, acronyms, and word boundary errors cause Twitter data to be unstructured and often noisy (Murshed et al., 2021). As a result, words may have been missed by code used and thus, quantitative data presented is more likely to be an underrepresentation of patterns in the original data set. Future research efforts should apply a cleansing/purification model to the tweet data to produce a more accurate reflection of themes in monkeypox-related digital content.

## Data Availability

All quantitative data files are available from the Harvard Dataverse database (doi:10.7910/DVN/CR7T5E).

https://dataverse.harvard.edu/dataset.xhtml?persistentId=doi:10.7910/DVN/CR7T5E

## Acknowledgements

I would like to thank Mary Tate for her invaluable mentorship throughout the research process. I would also like to thank N. Thakur for the open source tweet data used in the quantitative analysis.

## Notes

### Competing Interest Statement

The authors have declared no competing interest.

### Funding Statement

The author received no specific funding for this work.

